# Clinical Pilot of Bacterial Transcriptional Profiling as a Combined Genotypic and Phenotypic Antimicrobial Susceptibility Test

**DOI:** 10.1101/2024.07.10.24310021

**Authors:** EL Young, DJ Roach, MA Martinsen, GEG McGrath, NR Holbrook, HE Cho, EY Seyoum, VM Pierce, RP Bhattacharyya

## Abstract

**Brief Summary:** Exposure to antibiotics causes differential transcriptional signatures in susceptible vs. resistant bacteria. These differences can be leveraged to rapidly predict resistance profiles of *Escherichia coli* and *Klebsiella pneumoniae* in clinical positive blood cultures.

Antimicrobial resistance is a growing health threat, but standard methods for determining antibiotic susceptibility are slow and can delay optimal treatment, which is especially consequential in severe infections such as bacteremia. Novel approaches for rapid susceptibility profiling have emerged that characterize either bacterial response to antibiotics (phenotype) or detect specific resistance genes (genotype). GoPhAST-R is a novel assay, performed directly on positive blood cultures, that integrates rapid transcriptional response profiling with detection of key resistance gene transcripts, thereby providing simultaneous data on both phenotype and genotype. Here, we performed the first clinical pilot of GoPhAST-R on 42 positive blood cultures: 26 growing *Escherichia coli*, 15 growing *Klebsiella pneumoniae*, and 1 with both. An aliquot of each positive blood culture was exposed to 9 different antibiotics, lysed, then underwent rapid transcriptional profiling on the NanoString® platform; results were analyzed using an in-house susceptibility classification algorithm. GoPhAST-R achieved 95% overall agreement with standard antimicrobial susceptibility testing methods, with the highest agreement for beta-lactams (98%) and the lowest for fluoroquinolones (88%). Epidemic resistance genes including the extended spectrum beta-lactamase *bla*_CTX-M-15_ and the carbapenemase *bla*_KPC_ were also detected within the population. This study demonstrates the clinical feasibility of using transcriptional response profiling for rapid resistance determination, although further validation with larger and more diverse bacterial populations will be essential in future work. GoPhAST-R represents a promising new approach for rapid and comprehensive antibiotic susceptibility testing in clinical settings.

## INTRODUCTION

Infections due to antimicrobial resistant (AMR) bacteria are a major cause of global mortality, accounting for 1.27 million deaths in 2019 alone^1^, and result in increased length of hospital stays^2^, higher healthcare costs^3^, and higher mortality^2^. Delays in appropriate antimicrobial therapy, often due to bacterial resistance to empiric antibiotic selection, directly correlate with increased in-hospital mortality^4,5^. This can push clinicians toward excessively broad spectrum therapies that may result in worse patient outcomes^6^ and the development of resistance^7^, which underscores the broad potential benefits of rapid antimicrobial susceptibility testing (AST)^8,9,10^.

The gold standard for AST involves growing isolates in the presence of an antibiotic to determine the lowest concentration that inhibits bacterial growth^11^. While reliable, growth-based AST can be time-consuming, taking up to 72 hours from the time of sample collection to the final susceptibility profile. A growing number of alternative, rapid AST methods address this challenge in two broad ways: genotypic and phenotypic assays. Genotypic approaches directly identify specific genes or mutations known to confer resistance. This approach relies on detecting a limited subset of genes, however, and so is unable to detect novel or complex resistance mechanisms, especially in gram-negative pathogens^12^. Phenotypic assays, conversely, assess bacterial response to antibiotics based on different cellular properties, such as growth^13^, metabolic activity^14^, or bacterial motility^15,16^. Although generally applicable across resistance mechanisms, this approach does not provide information about bacterial genotypes, potentially omitting key data that could inform antibiotic selection^17^ and epidemiologic inquiries^18,19^.

In recent years, our group developed a method that combines both genotypic and phenotypic information into a single, rapid AST assay termed <u>G</u>en<u>o</u>typic and <u>Ph</u>enotypic <u>AST</u> through <u>R</u>NA detection (Go-PhAST-R)^20^. Go-PhAST-R uses NanoString® RNA hybridization chemistry^21^ to quantify multiple bacterial transcripts from crude lysate samples and infer resistance patterns. To do so, it leverages the marked differences in gene expression profiles of susceptible isolates compared to species-matched resistant isolates when exposed to antibiotics: resistant isolates are relatively unperturbed, whereas susceptible isolates, physiologically distressed and dying or growth-arrested, demonstrate large transcriptional changes in response to the antibiotic. We previously showed that a small subset of genes undergo large, predictable expression changes upon exposure to a class of antibiotics^22^, such that the change in expression of these marker genes reflects phenotypic antibiotic susceptibility, independent of resistance mechanism. Using this principle, we designed and validated GoPhAST-R probesets to classify susceptibility of *Escherichia coli* and *Klebsiella pneumoniae* to aminoglycosides, fluoroquinolones, and beta-lactams^22^. Additionally, transcripts from high-risk resistance genes, such as extended spectrum beta-lactamases (ESBLs) and carbapenemases, were simultaneously interrogated to identify high-risk genotypes of epidemiologic relevance. In prior work done on blood cultures spiked with laboratory strains with pre-specified resistance patterns, Go-PhAST-R achieved 94-99% accuracy, required minimal technical expertise and hands-on time, and returned results as soon as <4 hours after a positive blood culture^20^.

In this work, we implement this assay in a clinical setting for the first time, testing 42 blood cultures that grew *E. coli* or *K. pneumoniae* from the clinical microbiology laboratory at Massachusetts General Hospital (MGH). Transcriptional responses to antibiotic exposure clustered by susceptibility classification (i.e., susceptible strains and resistant strains exhibited distinct transcriptional responses), with 95% overall categorical agreement to standard clinical testing. These results are the first demonstration in a clinical setting of an assay based on the new paradigm of using bacterial transcriptional responses to predict antibiotic susceptibility.

## METHODS

### Bacterial Collection and Routine Culture Methods

A total of 66 unique positive blood cultures with gram-negative rods on Gram stain were collected; only one sample per patient was included. At the time a blood culture bottle signaled positive on the BACTEC FX system (Becton Dickinson, Sparks, MD), a 1mL aliquot was taken for processing as below. The remainder was carried forward for routine clinical processing, including subculture to solid media followed by colony identification via MALDI-TOF mass spectrometry (VITEK MS, version 3.2 in vitro diagnostic Knowledge Base, bioMérieux, Durham, NC) and antimicrobial susceptibility testing (AST) using the VITEK 2 AST-GN81 Gram Negative Susceptibility Card (bioMérieux), clinically validated for use with Clinical and Laboratory Standards Institute (CLSI) breakpoints published in M100-Ed31^23^. Aztreonam and cefazolin were tested by the CLSI disk diffusion method^24^ due to limitations of the VITEK 2 AST card; aztreonam was not included on the card, and the lowest concentration of cefazolin did not permit susceptible and intermediate minimum inhibitory concentrations (MICs) to be distinguished. Cefazolin susceptibility was further tested by broth microdilution^25^ in the research laboratory.

### Antibiotic treatment

As in prior implementations of GoPhAST-R^20^, a 1mL aliquot of each positive blood culture was first spun down at 100g for 10min to pellet red blood cells. The supernatant was then spun at 16,000g for 3min to pellet bacteria. The supernatant was removed, and the bacterial pellet resuspended in cation-adjusted Mueller-Hinton broth (CAMHB, ThermoFisher Scientific, Lenexa, KS) to a final volume of 500uL. Separate 45µl aliquots were added to pre-diluted 5 uL aliquots of cefazolin (CFZ), ceftriaxone (CRO), aztreonam (ATM), piperacillin-tazobactam (TZP), cefepime (FEP), ertapenem (ETP), meropenem (MEM), ciprofloxacin (CIP), levofloxacin (LVX), or gentamicin (GEN) to expose at CLSI breakpoint concentrations^23^ (**Table S1**). Bacteria were exposed at 37° C to each non-beta-lactam antibiotic for 60min, or 120min for beta-lactams given the slower response for this class^22^. In prior work, we found that the beta-lactam inoculum effect^26^ required exposure at standard CLSI-recommended inocula in order for the transcriptional response to reflect susceptibility^22^. Thus, beta-lactam treated samples and one untreated control were diluted 1:100 (targeting 5e5 cfu/mL) prior to incubation and plated at the time of antibiotic exposure to enumerate colony forming units to confirm. To assess baseline transcriptional signatures, two aliquots of each sample were each added to 5ul of CAMHB without antibiotics, and these untreated cultures were removed at either 60min (for comparison with aminoglycosides and fluoroquinolones) or 120min (for comparison with beta-lactams). Lysis buffer was added, and the samples were flash-frozen and stored at −80°C for subsequent batch processing.

### Lysis and Hybridization

Samples for which routine clinical testing identified *E. coli* or *K. pneumoniae* were thawed and lysed on the MP FastPrep™ bead beater^22^. Crude lysates were carried forward on the NanoString® platform per manufacturer’s protocols. RNA probe sets were designed as per prior work^20^. Samples were hybridized for 1 hour as per our prior protocol^20,22^, except for those treated with beta-lactams, which were hybridized for 16 hours to ensure sufficient signal from the lower initial inoculum.

### Quantification of Transcriptional Response and Genotyping

Data were partitioned by each antibiotic and species combination. Raw transcript counts were first normalized to NanoString® spike-in controls per manufacturer’s protocol, and then to housekeeping genes selected to have consistent expression to control for bacterial cellular density, as previously described^20^. To assess transcriptional response, we next calculated log_2_ fold change in normalized transcript counts between treated and untreated samples. To characterize resistance genotypes, probes were measured that targeted conserved regions of the carbapenemase genes *bla*_KPC_, *bla*_NDM_, *bla*_VIM_, *bla*_IMP_, and *bla*_OXA-48_ and the extended spectrum beta-lactamase gene *bla*_CTX-M-15_^20^. If any of these probes were detected above background, the gene was considered present.

### Model Construction and Resistance Prediction

To represent the entire transcriptional response profiles in a single quantitative metric, we used a method of one-dimensional projection called squared projected distance (SPD)^27^ to summarize each sample’s responsiveness in comparison to reference data from samples with known MICs. In brief, centroids representing the average transcriptional response of known susceptible and resistant isolates are computed for each antibiotic and species combination from highly susceptible and resistant strains from a reference set^22^. Distance from the susceptible centroid is calculated for each new transcriptional profile and scaled by the distance between the sensitive and resistant centroids; thus, an SPD of 0 represents a response similar to that of the control set of susceptible strains, whereas an SPD of 1 represents a resistant-like response.

We used support vector machine (SVM) modeling for each species and drug class as a tool to report the degree to which our susceptible and resistant isolates separate from each other by their SPD values. SVM is an unbiased, algorithmic method of identifying natural separation in data that minimizes overfitting by maximizing distance between boundaries and each class of data^28^. We implemented SVM models using the e1071 package^29^ on a dataset that included both the current samples as well as a larger collection with a higher proportion of resistant isolates from prior work^22^, using clinical susceptibility classifications as “ground truth”. To minimize the most clinically important discrepant classifications, we set the model to penalize very major discrepancies (VMDs) (R strain misclassified as S) twice as much as major discrepancies (S strain misclassified as R). For the genotypic portion of the assay, we incorporated whether resistance gene of a given class were detected, and if so, we considered the isolate to be resistant to the relevant antibiotic regardless of the SVM prediction (**Supplemental Fig. S1**). While the assay was run for cefazolin, the results were excluded from the final SVM model because (a) the CLSI breakpoint concentration falls within the MIC distribution of wild-type strains^30^, which in practice leads to classification challenges, exemplified in our collection by frequent discordance between susceptible and intermediate classifications for samples tested by disk diffusion and broth microdilution methods (**Supplemental Fig. S2**); and (b) cefazolin is not a recommended first-line therapy for gram-negative bacteremia, which was the focus of this study.

## RESULTS

### Bacterial collection and clinical AST determination

A total of 66 positive blood cultures with gram-negative rods on Gram stain were exposed in real-time in the clinical microbiology laboratory to the selected antimicrobials (Supplemental Table S1; see Methods). Of these 66 cultures, 26 were subsequently identified as *E. coli* and 15 as *K. pneumoniae*, for which we had previously designed probes for transcriptional susceptibility testing to beta-lactams, fluoroquinolones, and aminoglycosides^22^. In one case (sample 17), a blood culture was identified as having both species and was carried forward. The other 24 samples included species for which we do not have transcriptional probes and thus were not processed further. Routine clinical testing of these isolates found relatively low levels of resistance, with the highest rate of non-susceptibility found for ciprofloxacin at 26% (7/27) in *E. coli* and 19% (3/16) in *K. pneumoniae*, and the lowest rate seen for the carbapenems at 0% (0/27) and 6.3% (1/16) respectively (**Table 1**). While intermediate MICs were rare in the collection, we found 4 instances of intermediate MICs to levofloxacin in *E. coli* and 2 in *K. pneumoniae*.

**Table 1:** Isolates tabulated by their susceptibility profiles per standard testing in the clinical microbiology laboratory.

| Antibiotic | *E. coli* |  |  | *K. pneumoniae* |  |  |
| --- | --- | --- | --- | --- | --- | --- |
|  | S | I | R | S | I | R |
| Ceftriaxone (CRO) | 24 | 0 | 3 | 12 | 0 | 4 |
| Aztreonam (ATM) | 24 | 1 | 2 | 12 | 0 | 4 |
| Piperacillin<br>Tazobactam (TZP) | 27 | 0 | 0 | 14 | 1 | 1 |
| Cefepime (FEP) | 25 | 1 | 1 | 14 | 0 | 2 |
| Ertapenem (ETP) | 27 | 0 | 0 | 15 | 0 | 1 |
| Meropenem (MEM) | 27 | 0 | 0 | 15 | 0 | 1 |
| Ciprofloxacin (CIP) | 20 | 0 | 7 | 12 | 1 | 3 |
| Levofloxacin (LVX) | 17 | 4 | 6 | 12 | 2 | 2 |
| Gentamicin (GEN) | 23 | 0 | 4 | 14 | 1 | 1 |

### Transcriptional Response to Drug Treatment and Resistance Genotyping

To assess susceptibility from transcriptional profile, we measured levels of 10-13 transcripts per each drug class and calculated change in normalized expression between treated and untreated states^20^ (**Figure 1A**). While strain variation is present across drug treatments, *E. coli* and *K. pneumoniae* samples predominantly demonstrated one of two transcriptional profiles: susceptible isolates exhibited large perturbations in transcriptional responses upon antibiotic exposure, whereas resistant isolates demonstrated very little to no perturbation. When grouped by drug and species, transcripts from susceptible isolates were perturbed by an absolute log_2_-fold value of 1.0 to 4.0 averaged across all responsive genes, with most drug-species pairs falling between 1.5 and 3.0; while for resistant samples these values ranged from 0.2 to 1.8, with all but one falling below 0.9 (a carbapenem-resistant *K. pneumoniae*, see below) (**Figure 1B**). While uncommon, samples with intermediate MICs showed a wider average perturbation range of between 0.2 and 3.8 absolute log_2_-fold change.

**Figure 1.**
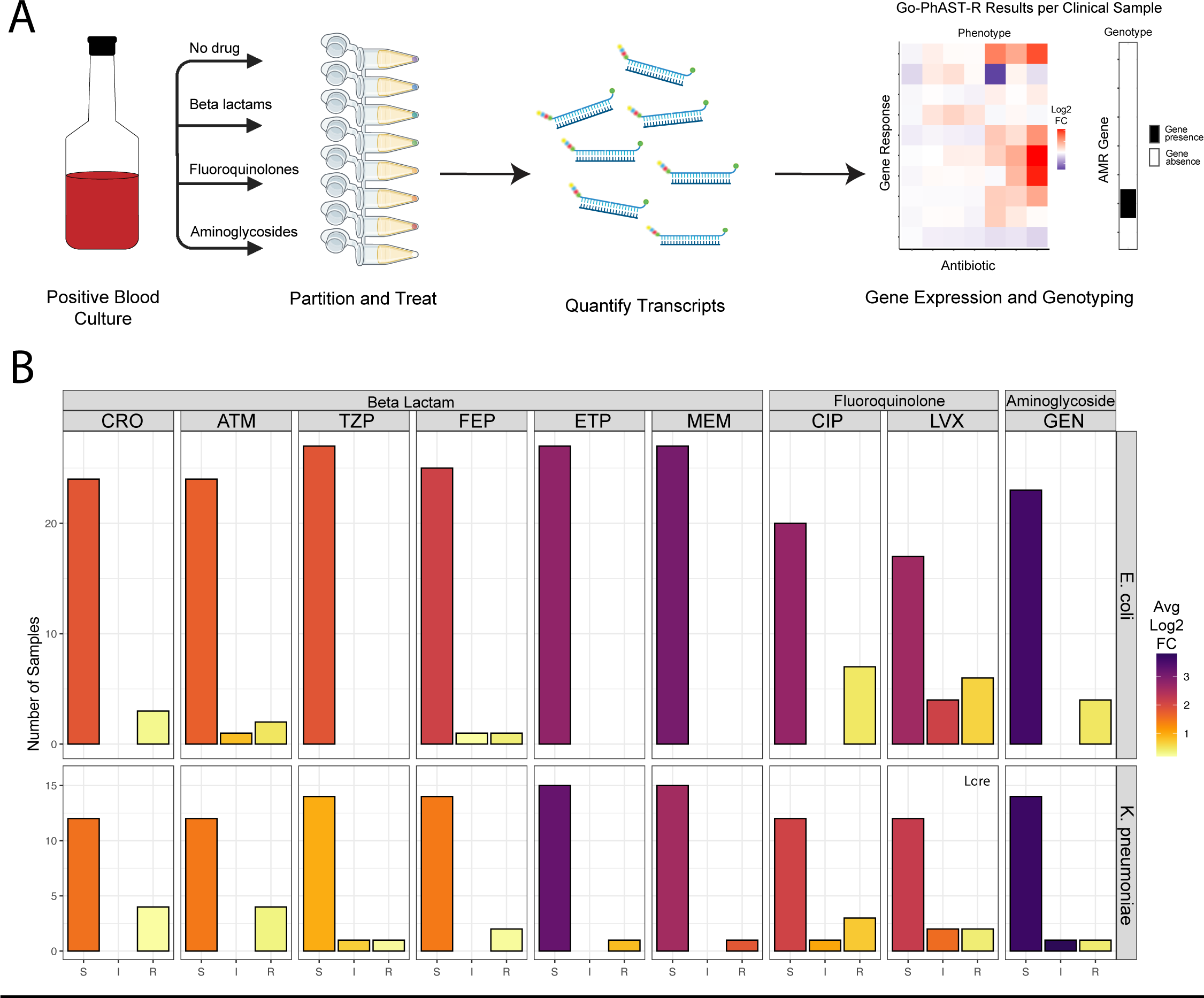
Assay Workflow and Summary of Transcriptional Profiles in Study Population. Transcriptional response to antibiotic exposure differs by susceptibility classification. (A.) Schematic workflow for an individual sample. (B) Clinical isolates show predictable transcriptional response to antibiotic treatment based on susceptibility profile. Absolute value of log2 fold-change of all transcript counts of genes targeted by the probeset upon antibiotic exposure averaged across isolates, subset by antibiotic and grouped according to clinical resistance profile (susceptible, intermediate, and resistant) and species (*E. coli* and *K. pneumoniae*). Antibiotics are abbreviated as per Table 1 and grouped by class. The color of each bar represents the average log2 fold change in transcript counts, while the height of each bar corresponds to the number of samples with that susceptibility profile for each antibiotic.

After condensing probe responses into squared projected distance (SPD)^27^ (**Figure 2A** and **2B, Supplemental Fig. S3**), a metric we previously devised to represent transcriptional response to antibiotic (see Methods), samples categorized as susceptible by clinical AST had a mean SPD value of 0.027 (median 0.0, interquartile range [IQR] ±0.09), while resistant samples had a mean SPD of 0.975 (median 1.0, IQR ±0.31). Samples with intermediate susceptibilities had a mean of 0.23 (median 0.1, IQR ±0.184) (**Figure 2C**).

**Figure 2:**
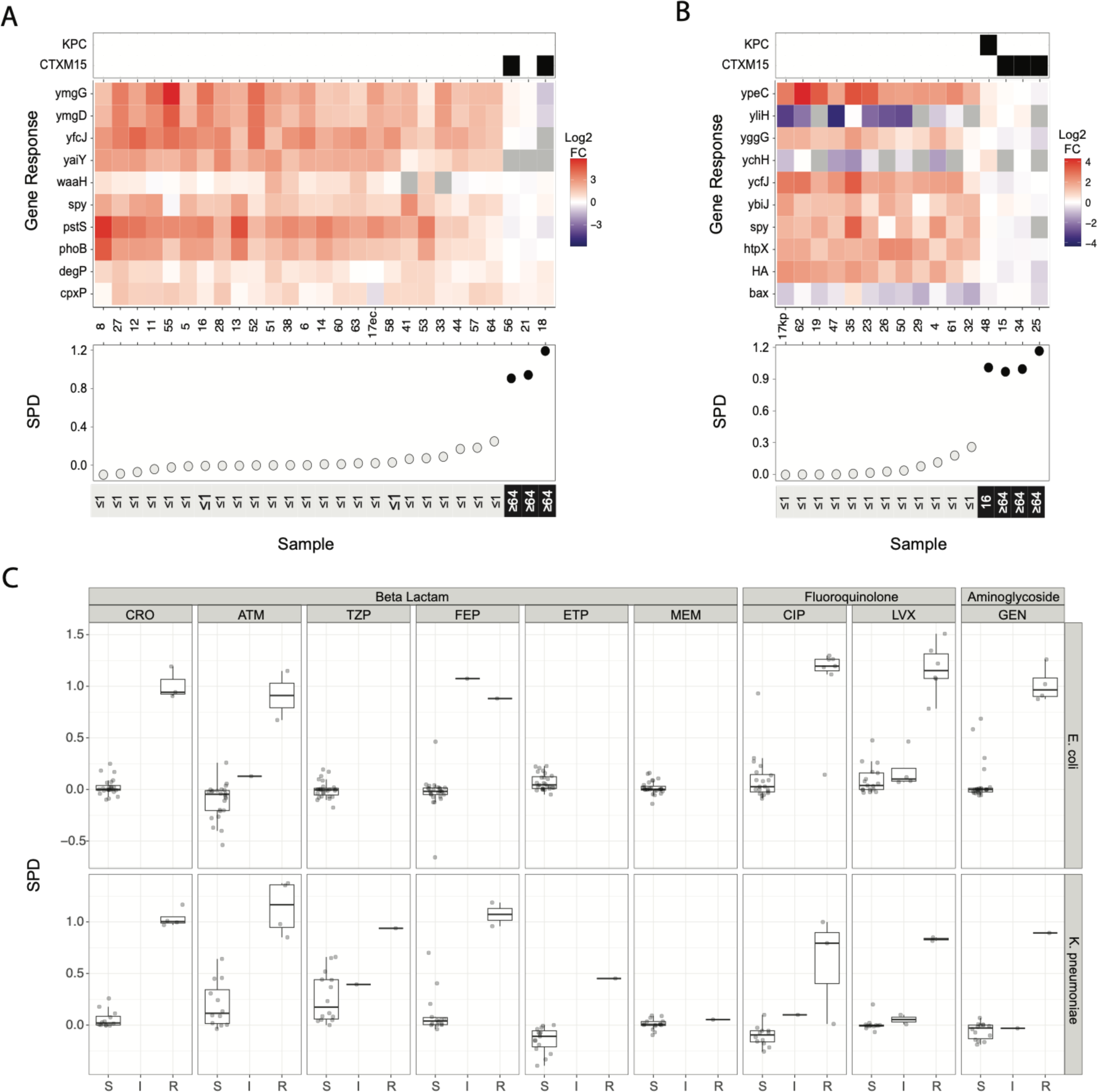
SPD Calculation from Transcriptional Response and Distribution. Transcriptional profile is distilled into a measure of treatment response with SPD, a single-value summary of the transcriptional response across the genes of interest (see Methods). (A-B). Heatmaps of the log2 fold change in transcriptional response of each (A) *E. coli* or (B) *K. pneumoniae* isolate after exposure to ceftriaxone. Each row corresponds to a different gene of interest, while each column is a different blood culture sample. KPC and CTX-M-15 genotypes are shown above the heatmap, while sample numbers, SPD values, and MICs (µg/mL) are shown below. (C) Box-and-whisker plot of SPD values for each species and antibiotic combination, grouped by clinical susceptibility profile. Antibiotic abbreviations are as listed in Table 1.

In total, our genotypic probes detected *bla*_CTX-M-15_ in 2 *E. coli* isolates and 3 *K. pneumoniae* isolates, and a single carbapenemase gene, a *bla*_KPC_ found in sample 48, a *K. pneumoniae* that displayed a transcriptional response to both ertapenem and meropenem despite harboring this resistance gene (**Supplementary Fig S4**).

### Susceptibility Predictions

We used a support vector machine (SVM) model to find the natural separation in the SPD values that best delineated susceptible and non-susceptible isolates (per clinical AST) and incorporated this into an algorithm for resistance classification (see Methods and **Supplementary Fig. S1**). We had an overall categorical agreement of 95% (368/387 pairwise drug comparisons) with standard susceptibility profiling (**Figure 3**). In total, there was a 5% very major discrepancy (VMD) rate (2/42 resistant isolates), a 2% major discrepancy (MD) rate (7/334 susceptible), and a 3% minor discrepancy rate (**Supplementary Table S2**). The best-performing grouping was the beta-lactam class in *E. coli*, with 99% overall categorical agreement and zero VMDs. The worst performing combination was in the fluoroquinolone class, with categorical agreement rates of 89% for *E. coli* and 88% for *K. pneumoniae* (**Supplementary Table S2**), and VMDs of 8% and 20%, respectively, representing one VMD in each species, with 12/13 correct resistance predictions for *E. coli* and 4/5 correct predictions for *K. pneumoniae*. Both misclassifications were in ciprofloxacin for isolates with VITEK 2 MICs at the resistant clinical breakpoint <u>(**Figure S3**)</u>.

**Figure 3:**
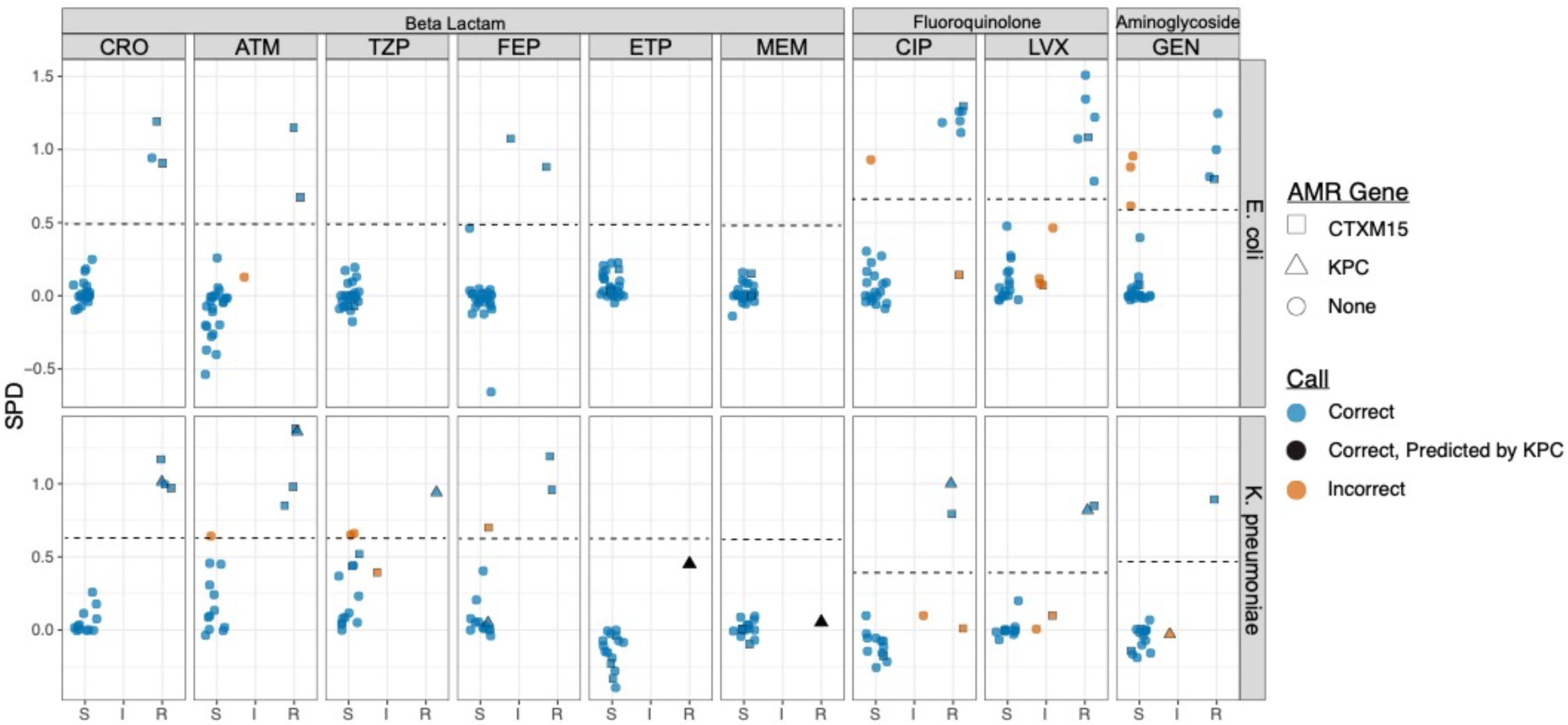
Predictions of Drug Susceptibility by SVM Modeling and Genotype. SVM thresholds for each drug class and bacterial species are shown by the dotted line. SVM is a model to find the optimal separation between two classes of data (see Methods). Each point corresponds to an SPD value (y-axis) and is shaped according to gene content and colored according to whether the SVM algorithm made the correct resistance assignment. Samples are grouped according to clinical susceptibility classification (x-axis).

Unexpectedly, we obtained one mixed culture of *E. coli* and *K. pneumoniae* during our collection (sample 17). Since each NanoString probe is species-specific in its reactivity pattern, we reasoned that we might see species-specific transcriptional responses for each species without subculture, so we carried the sample through gene expression profiling and analysis, using both *E. coli* and *K. pneumoniae* probesets for this sample (**Figure S3**, samples 17ec and 17kp). 18 of 20 SPD-based predictions for these two strains were in categorical agreement with clinical AST testing, with discrepancies only in fluoroquinolone susceptibility of the *E. coli* strain (one major discrepancy and one minor discrepancy for ciprofloxacin and levofloxacin, respectively; **Supplementary Table S3**). When this isolate was regrown and tested in laboratory monoculture, it demonstrated the expected susceptible transcription profile, with its SPD dropping from 0.93 to 0.15 (**Figure S5**). The *K. pneumoniae* in the mixed culture, by contrast, had perfect agreement with clinical AST results performed on the subcultured isolate (**Supplementary Table S3**).

## DISCUSSION

Here, we present the first pilot implementation of a transcription-based AST assay performed directly on positive blood cultures from clinical samples. This novel approach utilizes a single platform to concurrently assess bacterial phenotypic response to antimicrobials and identify high-risk AMR resistance genes in an integrated assay, culminating in a susceptibility prediction. The assay demonstrated strong overall concordance with routine AST methods, with some variation observed across antibiotic classes. Notably, beta-lactam antibiotics achieved 98% categorical agreement with standard clinical testing for six commonly used beta-lactams across both species and had no very major discrepancies (VMDs). Gentamicin also exhibited no VMDs in either species. However, two *E. coli* isolates classified as susceptible by standard testing were misclassified as resistant by the assay (major discrepancies) and one intermediate *K. pneumoniae* isolate was misclassified as resistant (minor discrepancy). Fluoroquinolones displayed the least favorable performance, with VMD rates of 8% (1/13) and 20% (1/5) in *E. coli* and *K. pneumoniae*, respectively, which represented the only VMDs across our entire sample set.

We examined each case in which the transcriptional response differed from expectation based on routine clinical susceptibility classification. In total, we observed four instances of resistant samples transcriptionally responding to treatment, which generally occurred in samples with MICs near the breakpoints (**Supplemental Fig. S3**). However, in two of these instances, the correct susceptibility prediction was made through genotypic detection. In these strain and antibiotic pairings, the *K. pneumoniae* isolate expressing *bla*_KPC_ (#48) showed considerable perturbations in the presence of both meropenem and ertapenem. While detecting *bla*_KPC_ is sufficient to eliminate carbapenem drugs as treatment options^31^, it has been noted previously that *bla*_KPC_-producing strains may display low MICs to carbapenems^32^, especially at low inocula^26^. In a different case, one blood culture (#17) was polymicrobial, growing both *E. coli* and *K. pneumoniae*. While the *K. pneumoniae* behaved as expected, the *E. coli* was susceptible to ciprofloxacin by MIC testing but did not demonstrate a strong transcriptional response, leading to a miscall as a resistant isolate by our model. The transcriptional response of this sample, however, looks qualitatively different than the “true” resistant profiles (**Supplemental Fig. S3**, ciprofloxacin panel, sample 17ec). On examination of the raw data from this sample, sample 17ec had low total transcript counts, likely introducing noise into the transcriptional signature, perhaps because *E. coli* was present at lower abundance than usual when the bottle signaled positive. As we have not previously included polymicrobial samples in the assay, further dedicated exploration will be needed to determine the optimal method for handling such samples. However, the ability in principle to simultaneously profile responses to multiple strains is a potentially appealing feature of this assay, as we observed 90% categorical agreement with standard AST testing across 20 phenotypic tests for these two isolates. The majority of the remaining errors were minor discrepancies in samples with MICs near CLSI breakpoints (**Fig S3**), which are inherently challenging classifications even in gold-standard AST methods^33,34^.

Our assay also captured genotypic information regarding the presence of key beta-lactamases, adding several useful features beyond phenotypic testing alone. First, as outbreaks of carbapenem-resistant bacteria may be caused by the spread of epidemic beta-lactamases^18^, knowledge about their presence has implications for infection control practices^35^. Second, a growing number of clinical decisions are determined by bacterial genotype. Current IDSA guidelines, for example, recommend carbapenems as first-line therapy for complicated infections caused by ESBL-producing organisms^17^. Finally, the emergence of novel beta-lactamase inhibitors with specificities for different carbapenemases underscores a role for gene identification to guide optimal antibiotic selection. Ceftazidime-avibactam, for instance, is effective against *bla*_KPC_ and *bla*_OXA-48_ carbapenemase-producing isolates but has no activity against metallo-beta-lactamases such as *bla*_NDM_^36^. As additional inhibitors to specific beta-lactamases are developed, the utility of defining genotype and phenotype together will likely grow in importance.

One limitation of our study is the low rates of bloodstream infections from resistant organisms within the cohort, resulting in a low number available for testing. However, we previously validated the assay on spike-in samples overrepresented for resistant isolates^20,22^. Second, the greatest discordance compared to standard testing occurred in those samples with MIC values near the clinical breakpoints, especially in the fluoroquinolone class. The inclusion of additional samples near breakpoints will be key to improving performance for this subset of isolates. Third, as CLSI clinical breakpoints evolve, the models would need to be retrained on updated guidelines to maintain the greatest accuracy. Fourth, our comparator method was not the reference standard of broth microdilution, but rather VITEK 2 (and disk diffusion in select cases), due to pragmatic considerations; however, this comparator reflects standard practices in many clinical microbiology laboratories, including our own. Most importantly, in this study we used SVM modeling to separate SPD values, and in order to have a sufficient number of resistant samples for model training, we combined data from prior work^22^ with the current population. This approach was necessary due to the low total numbers of resistant organisms, but as a result we do not have a fully independent validation set. We previously used a more sophisticated strategy using machine learning to train random forest models on the responses of each individual probe to create a robust classifier for meropenem, ciprofloxacin, and gentamicin susceptibility^20^. However, this approach requires larger training datasets, including a sizeable number of resistant isolates, to rigorously train and independently validate in a multistep process. Because each individual antibiotic within a class elicits a distinct magnitude of transcriptional response in each species (**Supplemental Fig. S2**), we could not use this original model without testing and retraining on hundreds of isolates, which was outside the scope of the current study. This additional training will be critical prior to clinical implementation and should improve classification accuracy. Finally, due to limited sensitivity of the stock NanoString nCounter Sprint detector system in our laboratory, we required a 16-hour hybridization time to detect signal from the lower inoculum samples used for beta-lactam testing. Pilot instruments under development offer improved RNA detection sensitivity with much shorter hybridization times^20^, which would bring beta-lactam testing times in line with other antibiotics.

In summary, in this study we perform the first clinical pilot using a diagnostic testing approach that simultaneously and rapidly detects bacterial phenotypic response to antibiotic exposure as well as epidemic AMR gene content. The synthesis of these complementary paradigms represents a promising step toward the next generation of clinical antimicrobial susceptibility testing. While further studies utilizing larger cohorts are necessary to refine the predictive algorithm and enhance its robustness, and automation of the process would be required before clinical utilization, this work establishes a framework for the continued development and ultimate clinical implementation of this novel transcriptional assay.

## Data Availability

All data generated and the code used for analysis are available on GitHub at https://github.com/broadinstitute/Go-PhAST-R_Clinical_Pilot/

## Acknowledgements

We thank the Massachusetts General Hospital Clinical Microbiology Laboratory for assistance with sample collection.

## Notes

### IRB approval

Clinical blood culture aliquots were collected under Mass General Brigham Institutional Review Board protocol 2015P002215.

### Financial support

This work was supported by the Massachusetts General Hospital Pilot Translational Research Grant (to RPB and VMP), the Broad Institute NextGen Award (to RPB), the National Institutes of Health (1R01AI153405, to RPB), and the John G. Bartlett Fellowship of the Antibiotic Resistance Leadership Group (T32AI007061-44, to DJR). The funders had no role in study design, data collection and interpretation, or the decision to submit the work for publication. The content is solely the responsibility of the authors and does not necessarily represent the official views of the National Institutes of Health.

### Conflicts of Interest

The authors declare no conflicts of interest.

## Notes

### Competing Interest Statement

The authors have declared no competing interest.

