## Supplemental Information for "Clinical Pilot of Bacterial Transcriptional Profiling as a Combined Genotypic and Phenotypic Antimicrobial Susceptibility Test"

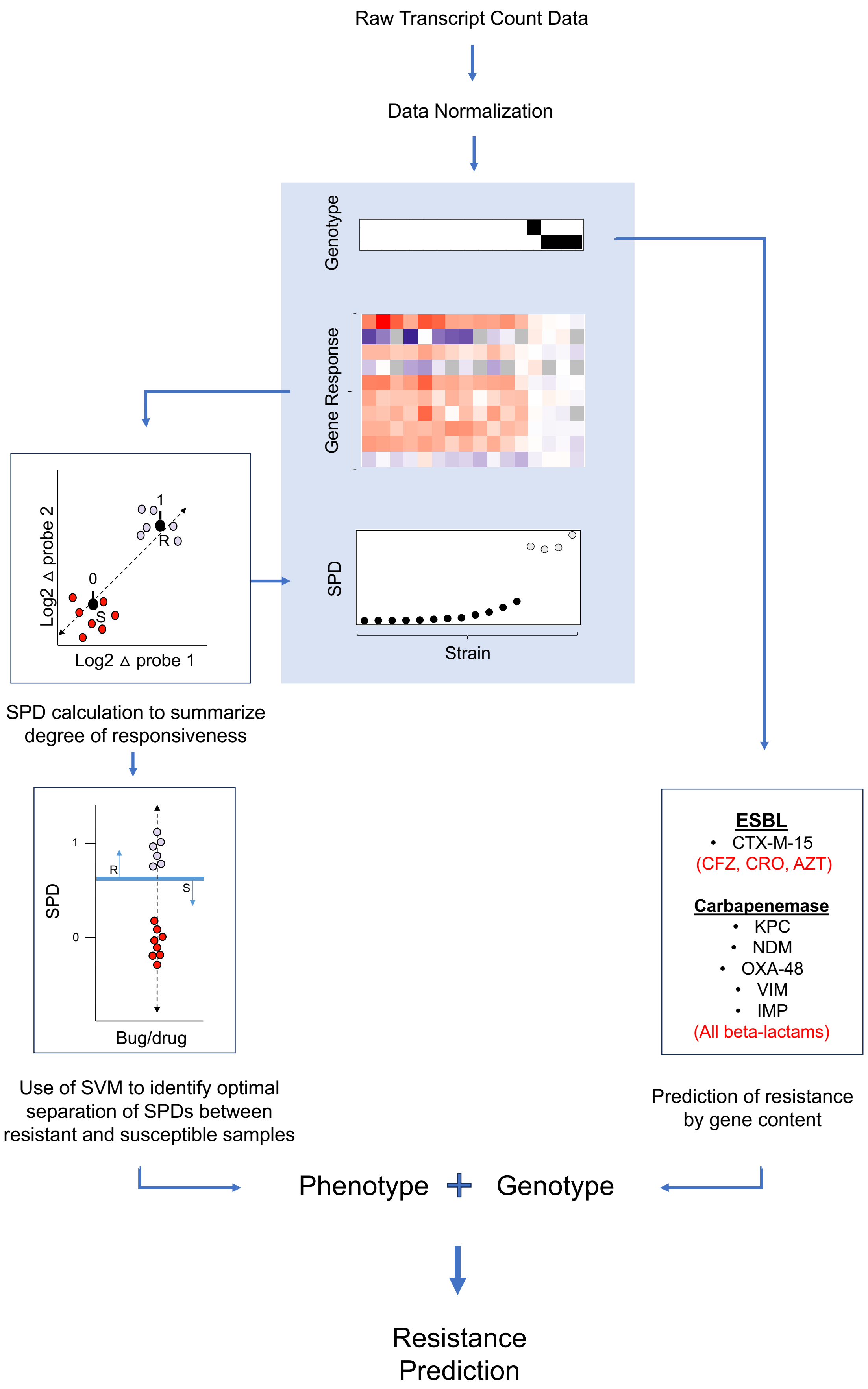


**Figure S1.** Analysis workflow for resistance prediction. The antibiotics to which a given gene would predict resistance are labeled in red.

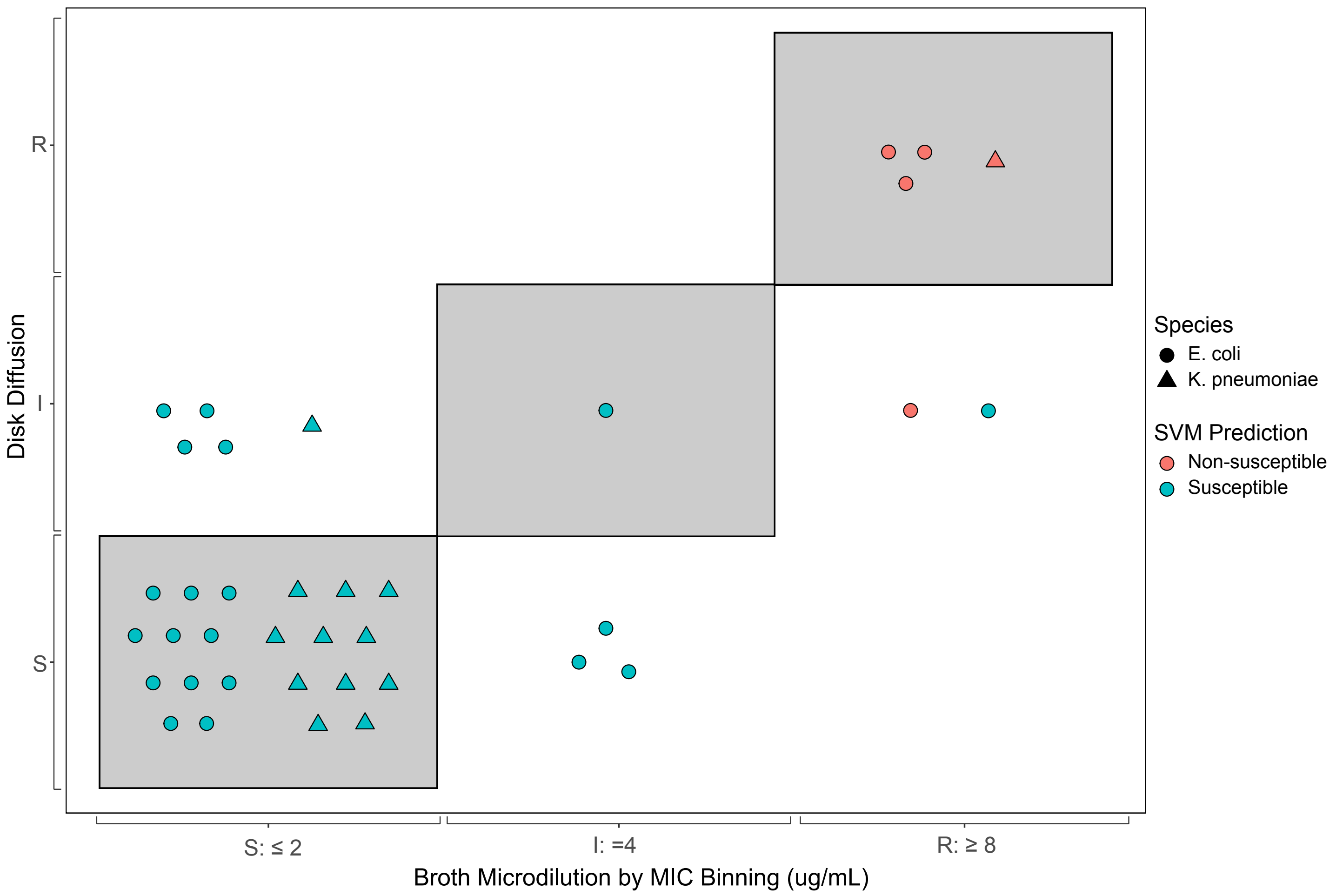


**Figure S2.** Discordant results for cefazolin between disk diffusion and broth microdilution. Results have been binned into the listed susceptibility categories. The plotted points have been jittered to separate them visually which does not represent variation in either MIC values or zones of inhibition; only the susceptibility class matters. As the VITEK 2 AST-GN81 card does not discriminate between susceptible and intermediate cefazolin MICs, isolates were tested by disk diffusion in the clinical microbiology laboratory, with results shown on the y-axis. Isolates were all subsequently tested by broth microdilution by the research laboratory, with results shown on the x-axis. In total we retested 37 isolates, including all 33 susceptible and intermediate isolates, which resulted in 10 discordant results between broth microdilution and disk diffusion (shown as isolates outside the grey boxes). The broth microdilution and disk diffusion susceptibility classifications corresponded at similar rates with expected transcriptional response profiles (concordance of 32/37 and 30/37, respectively). Given that the CLSI breakpoint concentration falls within the MIC distribution of wild-type strains (leading to challenges with classification of susceptible and intermediate MICs by any phenotypic AST), together with the fact that cefazolin is not a recommended first-line therapy for gram-negative bacteremia, it was removed from further inclusion in the study.

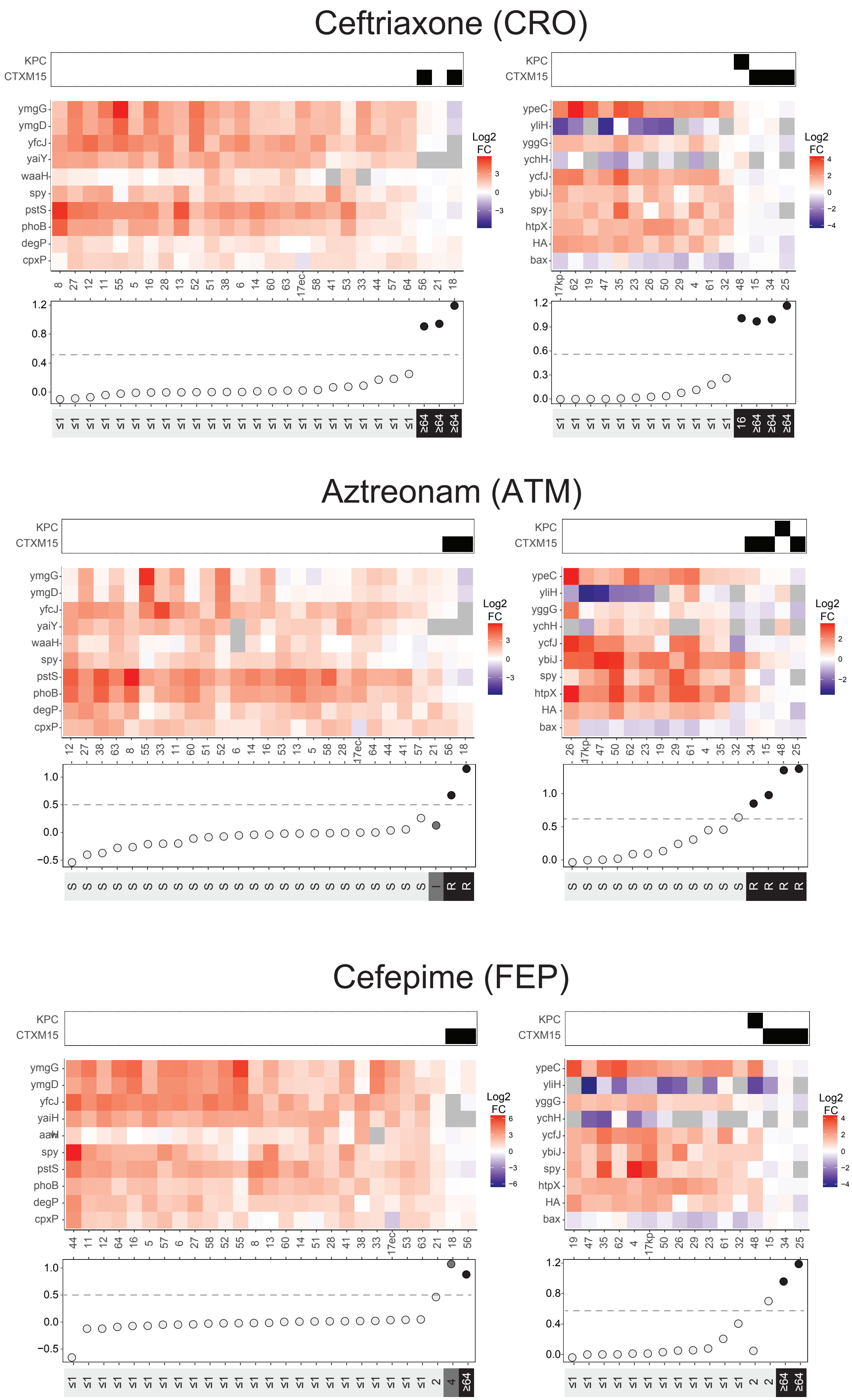

Ceftriaxone (CRO)
KPC
CTXM15
ymgG
ymgD
yfcJ
yaiY
waaH
spy
pstS
phoB
degP
cpxP
ypeC
yliH
yggG
ychH
ycfJ
ybiJ
htpX
HA
bax
Log2 FC
Aztreonam (ATM)
Cefepime (FEP)

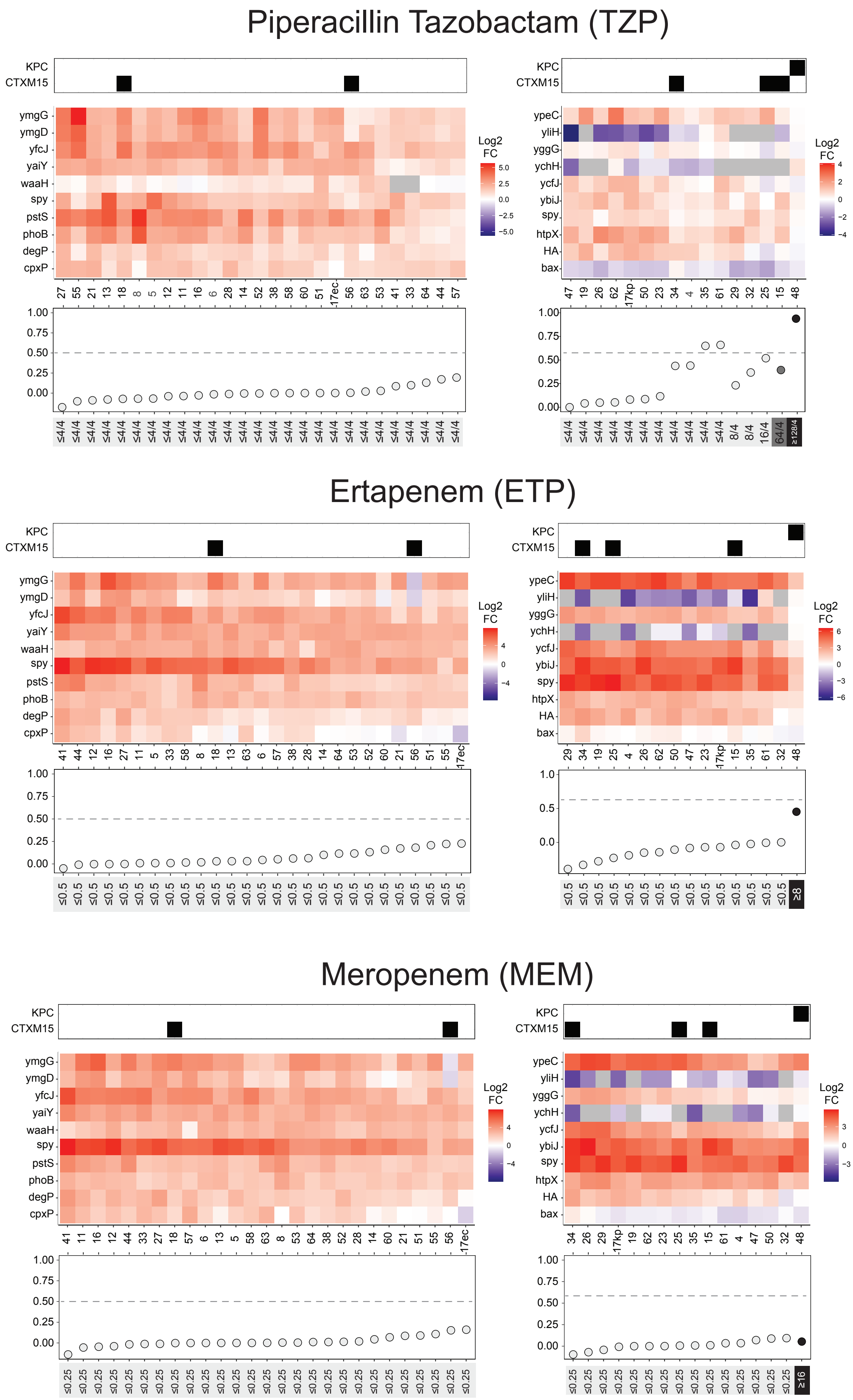
Piperacillin Tazobactam (TZP)
KPC
CTXM15
ymgG
ymgD
yfcJ
yaiY
waaH
spy
pstS
phoB
degP
cpxP
Log2 FC
5.0
2.5
0.0
−2.5
−5.0
ypeC
yliH
yggG
ychH
ycfJ
ybiJ
spy
htpX
HA
bax
Log2 FC
4
2
0
−2
−4
1.00
0.75
0.50
0.25
0.00
≤4/4
8/4
16/4
64/4
≥128/4
Ertapenem (ETP)
KPC
CTXM15
Log2 FC
4
0
−4
6
3
0
−3
−6
1.0
0.5
0.0
≤0.5
≥8
Meropenem (MEM)
KPC
CTXM15
Log2 FC
4
0
−4
3
0
−3
≤0.25
≥16

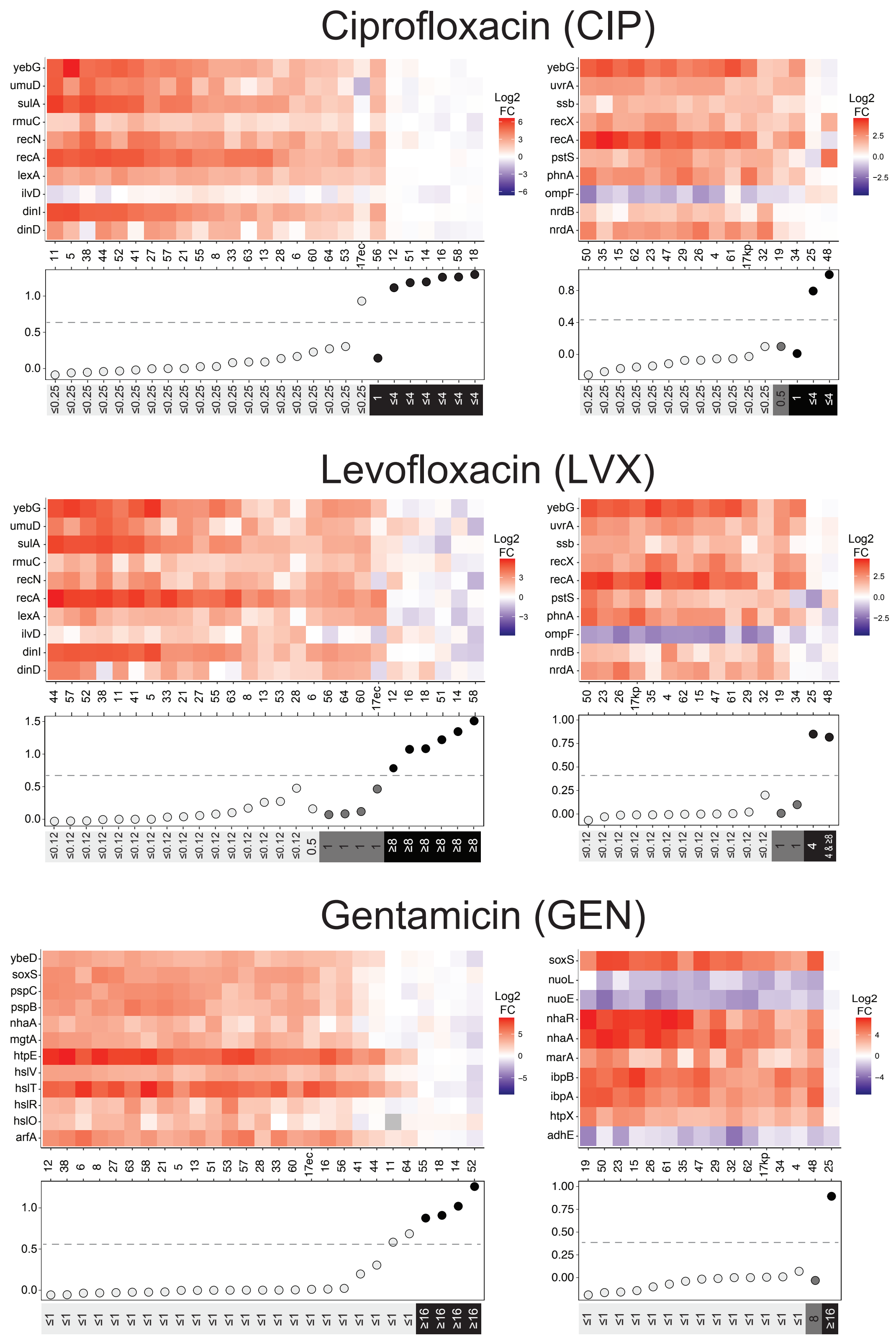


**Figure S3.** Heatmaps of the log2 fold change in transcriptional response of each *E. coli* (left) or *K. pneumoniae* (right) isolate after exposure to the listed antibiotic. Each row corresponds to a different gene of interest, while each column is a different blood culture sample. The KPC and CTX-M-15 genotypes are shown above the heatmap for the beta-lactam antibiotics, while the SPD values and MICs are shown below with the SVM thresholds delineated with the dotted lines. Sample 17 is the polymicrobial culture.

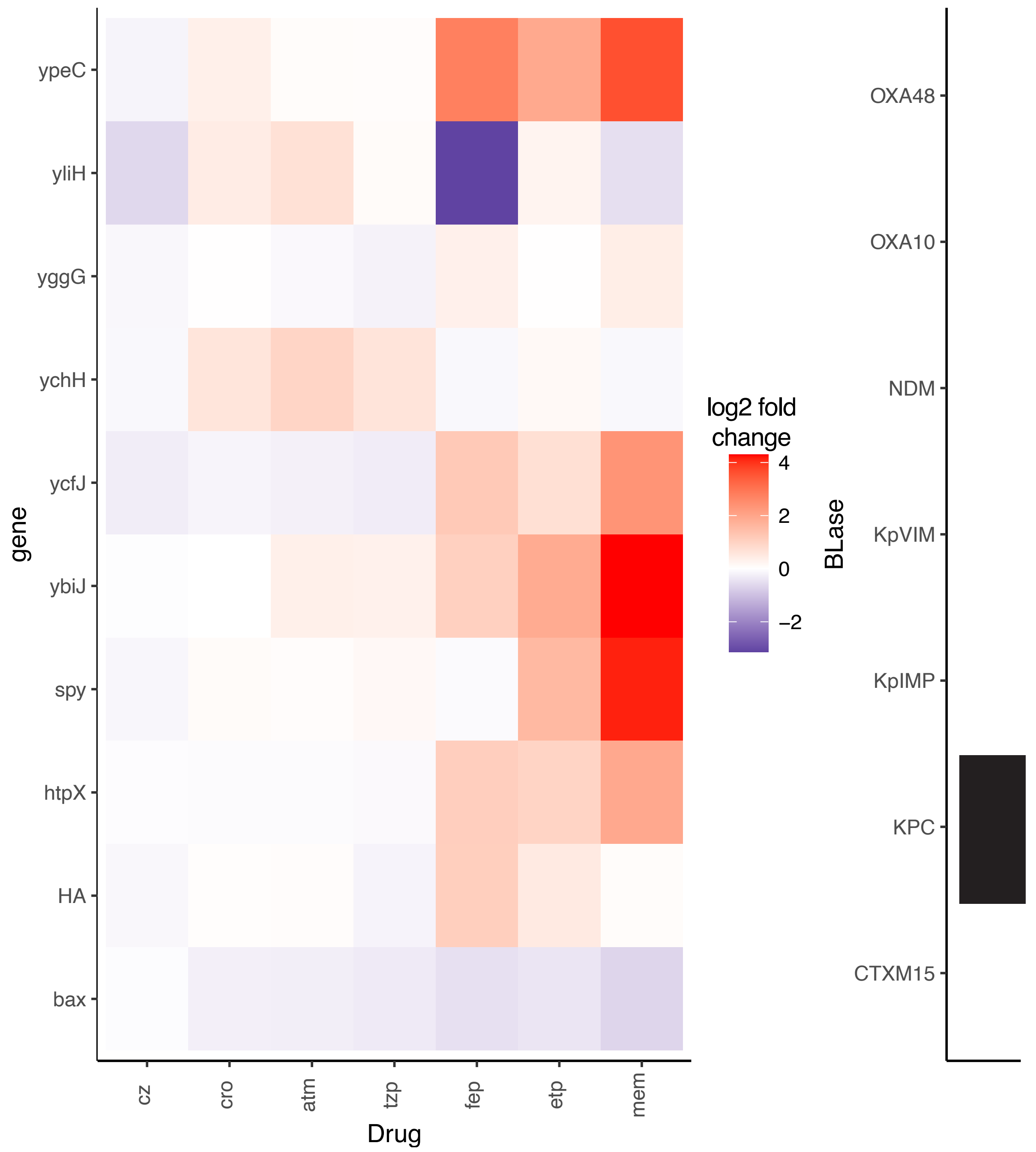


**Figure S4.** Beta-lactam restricted heatmap for carbapenem-resistant (by MIC) *K. pneumoniae* isolate #48, which was predicted to be carbapenem-susceptible by SVM due to the significant transcriptional response to these antibiotics but had KPC transcripts on genotyping.

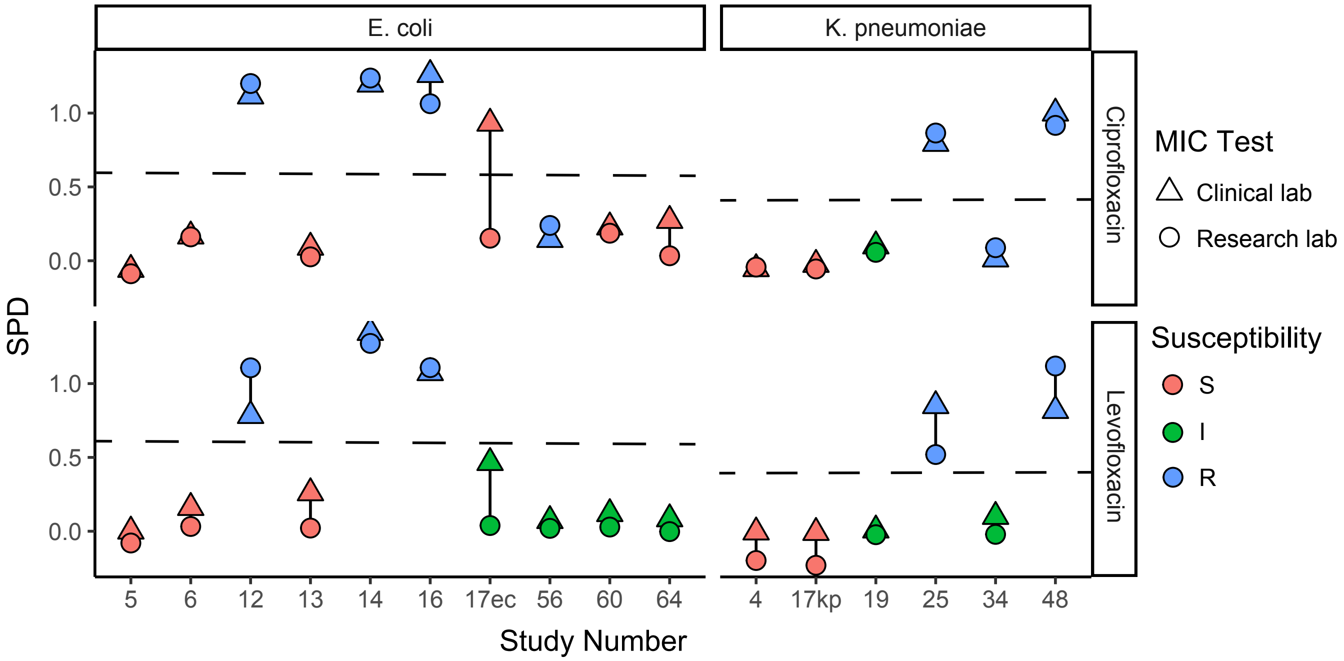


**Figure S5.** Comparison of clinical laboratory and research laboratory SPDs for fluoroquinolones SVM thresholds are demonstrated by the dotted line. A subset of 10 *E. coli* and 6 *K. pneumoniae* were re-run in the laboratory after regrowing in broth. The only case which demonstrated a large difference in SPD (resulting in a change in resistance prediction) was the *E. coli* isolate (17ec) from a blood culture sample that was polymicrobial. The *K. pneumoniae* isolate (17kp) did not demonstrate a significant shift in SPD.

| Drug | Dose (μg/mL) |
|---|---|
| Ciprofloxacin | 0.25 |
| Levofloxacin | 0.5 |
| Gentamicin | 4 |
| Cefazolin | 2 |
| Ceftriaxone | 1 |
| Cefepime | 2 |
| Aztreonam | 4 |
| Piperacillin-tazobactam | 16/4 |
| Ertapenem | 0.5 |
| Meropenem | 1 |

**Table S1:** Treatment doses for each drug in the study. Each sample was treated with breakpoint concentrations current at the time the study was conducted.

| | | Number Tested | Susceptible | Intermediate | Resistant | Very Major Disc. Rate | Major Disc. Rate | Minor Disc. Rate | Categorical Agreement |
|---|---|---|---|---|---|---|---|---|---|
| ***Eschericia coli*** | **Beta Lactam** | 162 | 154 | 2 | 6 | 0 | 0 | 0.006 | 0.99 |
| | **Fluoroquinolone** | 54 | 37 | 4 | 13 | 0.077 | 0.027 | 0.074 | 0.89 |
| | **Aminoglycoside** | 27 | 23 | 0 | 4 | 0 | 0.087 | 0 | 0.93 |
| ***Klebsiella pneumoniae*** | **Beta Lactam** | 96 | 82 | 1 | 13 | 0 | 0.049 | 0.01 | 0.95 |
| | **Fluoroquinolone** | 32 | 24 | 3 | 5 | 0.2 | 0 | 0.094 | 0.88 |
| | **Aminoglycoside** | 16 | 14 | 1 | 1 | 0 | 0 | 0.063 | 0.94 |
| | Total | 387 | 334 | 11 | 42 | 0.048 | 0.021 | 0.026 | 0.95 |

**Table S2:** Predictions of the SVM and KPC detection are tabulated by species and drug class, as well as cumulatively.

| | 17Ec | | | | 17Kp | | | |
|---|---|---|---|---|---|---|---|---|
| | SPD | Prediction | Clinical AST | Result | SPD | Prediction | Clinical AST | Result |
| **CFZ** | 0.27 | S | S | Correct | 0.04 | S | S | Correct |
| **CRO** | 0.02 | S | S | Correct | 0.00 | S | S | Correct |
| **ATM** | 0.00 | S | S | Correct | 0.00 | S | S | Correct |
| **TZP** | 0.00 | S | S | Correct | 0.08 | S | S | Correct |
| **FEP** | 0.04 | S | S | Correct | 0.01 | S | S | Correct |
| **ETP** | 0.23 | S | S | Correct | -0.07 | S | S | Correct |
| **MEM** | 0.16 | S | S | Correct | -0.01 | S | S | Correct |
| **CIP** | 0.93 | NS | S | Maj Discrepancy | -0.03 | S | S | Correct |
| **LVX** | 0.46 | S | I | Min Discrepancy | -0.01 | S | S | Correct |
| **GEN** | 0.01 | S | S | Correct | 0.00 | S | S | Correct |

**Table S3.** AST results of the polymicrobial sample with the *E. coli* isolate results on the left and the *K. pneumoniae* isolate results on the right.
